# The arrival and spread of SARS-CoV2 in Colombia

**DOI:** 10.1101/2020.06.11.20125799

**Authors:** Juan David Ramírez, Carolina Florez, Marina Muñoz, Carolina Hernández, Adriana Castillo, Sergio Gomez, Angelica Rico, Lisseth Pardo, Esther C. Barros, Sergio Castañeda, Nathalia Ballesteros, David Martínez, Laura Vega, Jesús E. Jaimes, Lissa Cruz-Saavedra, Giovanny Herrera, Luz H. Patiño, Aníbal A. Teherán, Ana S. Gonzalez-Reiche, Matthew M. Hernandez, Emilia Mia Sordillo, Viviana Simon, Harm van Bakel, Alberto Paniz-Mondolfi

## Abstract

We performed phylogenomic analysis of severe acute respiratory syndrome coronavirus-2 (SARS-CoV2) from 88 infected individuals across different regions of Colombia. Eleven different lineages were detected, suggesting multiple introduction events. Pangolin lineages B.1 and B.1.5 were the most frequent, with B.1 being associated with prior travel to high-risk areas.

**Article Summary Line:** We sequenced 88 genomes of SARS-CoV2 from Colombia finding 11 lineages and eight different introduction events

---

The number of severe acute respiratory syndrome coronavirus 2 (SARS-CoV2) infections is rapidly increasing throughout South America (SA). Colombia, the fifth largest country in SA is also the fifth country in number of confirmed new coronavirus disease-2019 (COVID-19) cases in the region by June 8^th^, 2020. Following identification of the first COVID-19 case in Colombia, on March 6, 2020, in a female traveler returning from Milan, Italy, the Colombian government implemented early control measures. An ongoing mandatory lockdown and travel ban/restriction was put in place on March 23, 2020, which included the closure of all the airports across the country. Although these containment measures certainly helped reduce the basic reproduction number (*R*_0_) from 4.8 to 2.2, they have been unable to fully limit SARS-CoV2 spread in Colombia [1].

Colombia’s geographic location makes it an important crossroad in the Andean region, attracting many travelers across SA and making it a particular vulnerable region with important implications for internal spread and dissemination to its multiple bordering countries. However, in-depth studies on the molecular epidemiology and SARS-CoV2 strains circulating in Colombia and elsewhere in SA are still lacking.

## The Study

Individuals meeting case-definition criteria established by the Colombian Ministry of Health and Social Protection were screened for SARS-CoV2 infection at different hospitals and healthcare centers in 16 of the 32 Departments of Colombia between March 31^st^ and May 1^st^ 2020 [1]. Molecular detection of SARS-CoV2 in clinical nasopharyngeal swab (NP-VTM) specimens was performed using the Berlin Charité protocol [1], with 88 positive cases selected for further study. Most of the SARS-CoV2 infected patients were identified in 4 (Andean, Caribbean, Pacific and Orinoco) of the 6 geographic regions of Colombia; particularly in the Departments of Valle del Cauca (35.9%), Cundinamarca (11.2%), Boyacá (10.1%), Antioquia (8.9%) and Huila (7.8%). Sociodemographic characteristics of the 88 SARS-CoV2-positive patients showed that the average age was 44 (ranging from 36-58), with 58% (n=51) male and 42% (n=37) female subjects. Different risk factors for exposure were identified, 12 (13.6%) patients were health care workers, 55 (62%) had close contact with infected patients and 23 (28.4%) had traveled to high-risk areas (Mostly European countries). On presentation 17 (19.3%) were asymptomatic, 71 (80.7%) were symptomatic and 26 (29.5%) required hospitalization. At presentation, the most common symptoms were respiratory (80.6%), fever (59%) and gastrointestinal symptoms (33%). Respiratory symptoms ranged from nonspecific influenza-like symptoms (dry cough and shortness of breath) to respiratory failure (5.7%). Twenty-nine (33%) patients had concurrent conditions such as diabetes, hypertension, COPD, asthma, cardiac failure and cancer.

To assess the genetic diversity and origins of SARS-CoV2 in Colombia, we sequenced and assembled viral genomes from total RNA extracted from NP-VTM clinical specimens, as described elsewhere [1]. Comparative genome analysis of our 88 cases and 3 previously reported Colombian cases was carried out relative to publicly available background data from 2,744 cases sampled from the GISAID EpiCoV database to obtain a full representation of global lineage diversity [1]. Lineage assignments were performed using the Phylogenetic Assignment of Named Global Outbreak LINeages tool ‘Pangolin’ [1]. Consensus viral sequences from each case were also submitted to GISAID (accessions: EPI_ISL_447734-EPI_ISL_447817).

The Maximum Likelihood (ML) reconstruction showed that the Colombian genomes have a close phylogenetic relation to a wide range of SARS-CoV2 strains across 11 different Pangolin lineages [1], with a predominance of B lineages all across the country (Fig. 1). We performed univariate analysis to determine whether certain lineages were associated with a health-care worker status, hospital exposure (including intensive care unit admission), or a travel history to high-risk areas. A significant association (p=0.033) was only found between infection with B.1 lineage and a travel history to high-risk areas. In conclusion, Pangolin lineage B.1 was associated with prior travel to high-risk areas.

**Fig 1.**
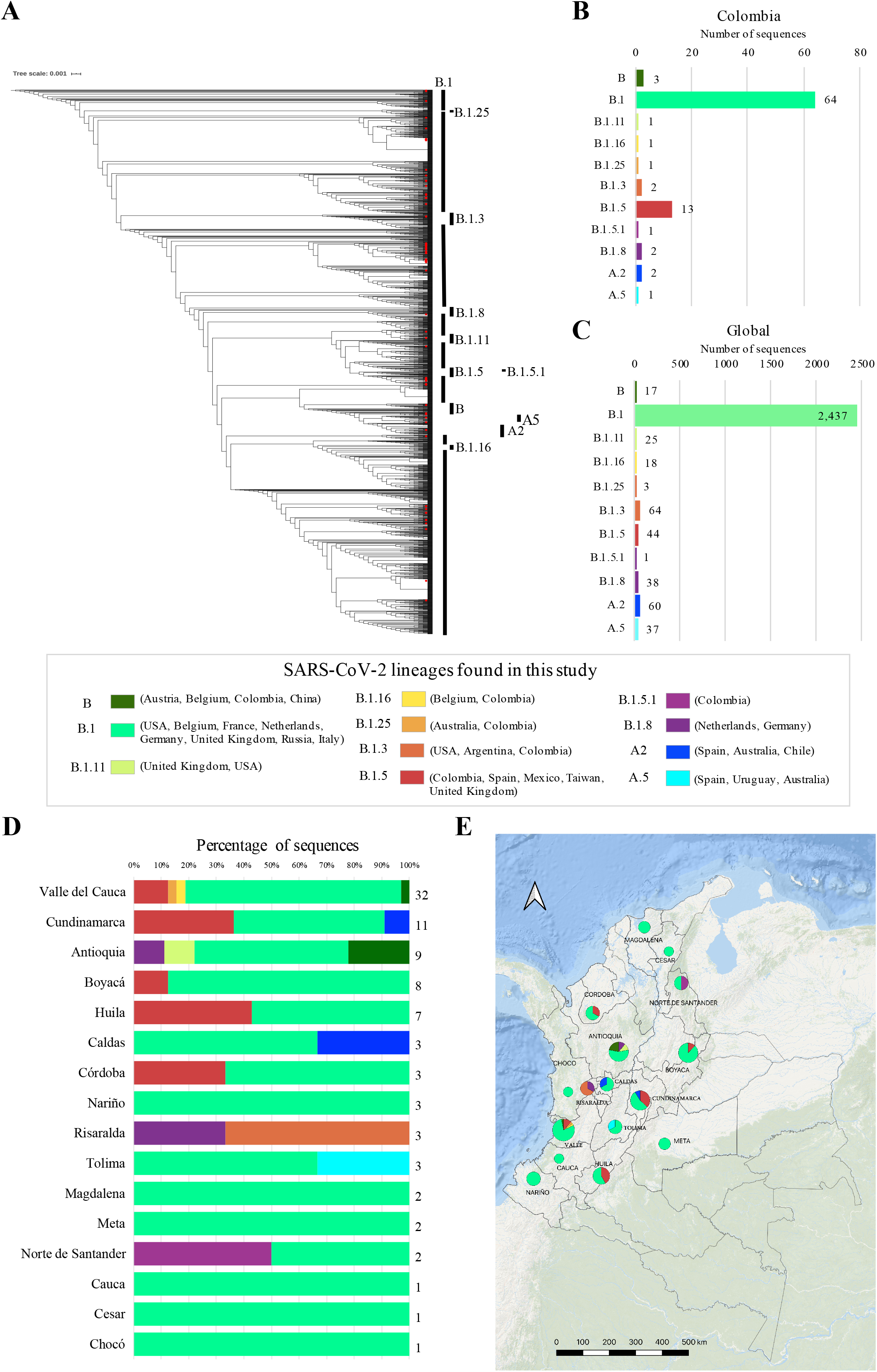
Distribution of several SARS-CoV2 lineages in Colombia. **A**. Maximum likelihood (ML) tree of 2,744 global background and 91 Colombian genomes (red dots; including 3 genomes previously deposited in GISAID), the tree was rooted with Pangolin coronavirus (MT084071.1). The pangolin nomenclature is used to show the eleven lineages detected in the country. **B**. Percentage of Colombian genomes assigned to a specific Pangolin lineage. **C**. Percentage of genomes from the global diversity (highlighting their geographical origin) assigned to the lineages described in Colombia using Pangolin nomenclature **C**. Percentage of genomes belonging to the different lineages by Department of Colombia, assumed as geographical regions according with the national administrative and political division. **D**. The geographical distribution of SARS-CoV2 lineages in Colombia using IQGIS.

We further constructed a time-scaled ML phylogeny using TreeTime and specimen collection date constraints [1]. At least nine potential introductions during the dispersion of SARS-CoV2 into the country were identified between January 20^th^ (CI95% Jan 18^th^-Jan 20^th^) to March 12^th^ (CI95% March 11^th^ −12^th^) (Fig. 2). This suggests that SARS-CoV2 may have been circulating cryptically in Colombia between late January and mid-March; up to two months before confirmation of the first official case of COVID-19 by the Colombian Ministry of Health and Social Protection.

**Fig 2.**
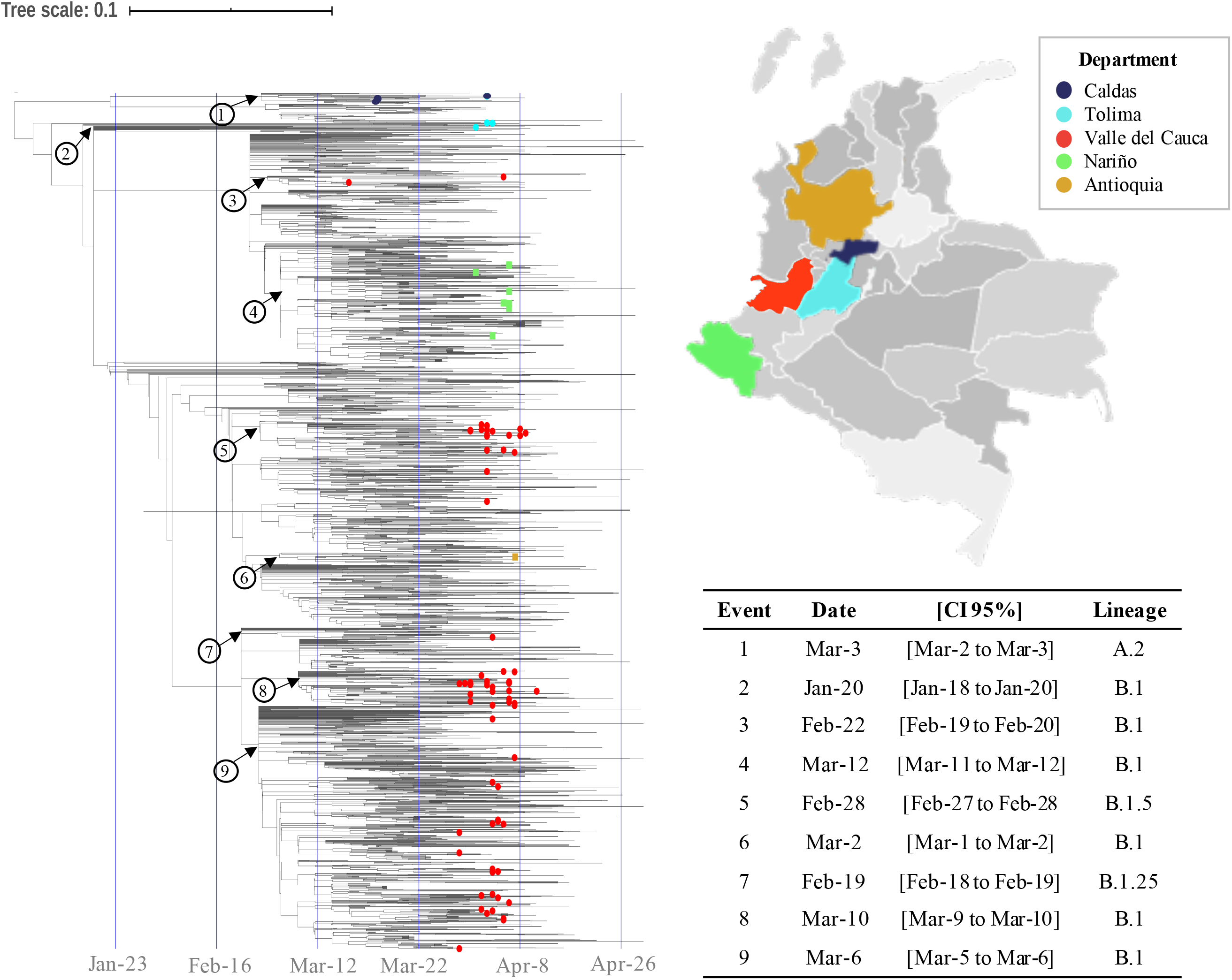
Multiple and early introductions of SARS-CoV2 lineages in Colombia. Time-scaled tree built in TreeTime from the trimmed whole genome alignment of the global background diversity (left). The colored dots indicate the 91 genomes encompassing the 11 lineages herein described. Dots are colored according to the introductions to specific geographical regions of Colombia shown on the right and labeled with numbers in the tree (Turquoise=Tolima, Red= Valle del Cauca, Yellow=Antioquia, Purple=Caldas and Green=Nariño). The nodes with the dates estimates are indicated with the blue arrows and the number of introductions with the numbers and time estimates in the table (right). The displayed time tree was inferred under a strict clock model with a fixed substitution rate of 0.8×10^−3^, based on previous rate value estimates [1]. TreeTime analyses were run for a total of 6 iterations and marginal date estimates of ancestral states are shown with 90% confidence intervals.

## Conclusions

Our data supports variable sources of introductions of the SARS-CoV2 into Colombia predominantly from Europe and North America (Fig. 1), as well as active transmission despite establishment of early containment measures prior to the date of the first case detected (Fig. 2). Three scenarios may explain such trend. First, infected travelers and migrants from countries already affected by the SARS-CoV2 likely entered Colombia prior to the travel ban and closure of Colombia’s borders (March 23^rd^, 2020). Second, many Colombian citizens with limited economic and/or social resources have been unable to comply with quarantine measures. Third, vast variations in ethnicity, climate and sociodemographic features (environmental heterogeneity) across Colombia may be influencing the presentation and spread of the virus.

Our findings indicate that B1 and B1.5 are currently the most widespread SARS-CoV-2 lineages in Colombia. Our data also suggest, based on the initial description of these lineages, that there have been mainly introductions from Europe and the USA, with additional introductions from China, Australia, Mexico, Chile and Uruguay (Fig. 1). This highlights the pivotal importance of genomic surveillance in epidemics and its direct implications on inferring dissemination patterns. A limitation of this initial study is that we received specimens by referral and did not systematically sample different regions of the country.

The arrival of SARS-CoV2 in SA poses particular challenges, as the virus now spreads across a region with diverse and complex geopolitical and sociocultural contexts. Marked poverty, urban and suburban overcrowding, scarce sanitary conditions as well as overwhelmed public health systems sharply contrast with the way SARS-CoV2 has impacted most of the industrialized countries of the world so far. Such conditions may negatively impact viral dynamics favoring transmission and long-term persistence. In fact, the WHO has recently stated that SA has now become the new epicenter of the global coronavirus pandemic, thus urging implementation of widespread population surveillance and reinforcing containment measures.

Future studies in Colombia and elsewhere in SA, including sequencing of viral genomes as the predicted epidemic peak approaches, and of contact cases and spread clusters, may help to better identify transmission routes and inform potential prevention measures. Our study supports the relevance of genomic surveillance and the critical need to establish coordinated efforts to generate genomic data in SA that will enable integrative analyses to uncover SARS-CoV-2 dynamics at the continental level.

## Data Availability

All data related with this manuscript is deposited on GISAID.

## Acknowledgments

We thank Dirección de Investigación e Innovación from Universidad del Rosario for funding this study.

## Author Bio

Juan David Ramírez, Ph.D. is associate professor at Universidad del Rosario in Bogotá, Colombia. His primary research interests are genomic epidemiology and, ecology and evolution of infectious diseases.

